# Renin-angiotensin-aldosterone system inhibitors and mortality in patients with hypertension hospitalized for COVID-19: a systematic review and meta-analysis

**DOI:** 10.1101/2020.05.21.20107003

**Authors:** Anna E. Ssentongo, Paddy Ssentongo, Emily S. Heilbrunn, Alain Lekoubou, Ping Du, Duanping Liao, John S. Oh, Vernon M. Chinchilli

## Abstract

**Objective:** The association between renin-angiotensin-aldosterone (RAAS) inhibitors and Coronavirus diseases 2019 (COVID-19) mortality is unclear. We aimed to explore the association of RAAS inhibitors, including angiotensin-converting inhibitors (ACEi) and angiotensin II receptor blockers (ARBs) with COVID-19 mortality in patients with hypertension.

**Methods:** MEDLINE, SCOPUS, OVID, and Cochrane Library were searched for the period of January 1, 2020 to May 20, 2020. Studies reporting the association of RAAS inhibitors (ACEi and ARBs) and mortality in patients with hypertension, hospitalized for COVID-19 were extracted. Two reviewers independently extracted appropriate data of interest and assessed the risk of bias. All analyses were performed using random-effects models on log-transformed risk ratio estimates, and heterogeneity was quantified.

**Results:** Data were collected on 2,065,805 individuals (mean age, 58.73 years; 53.4% male). Patients with hypertension taking RAAS inhibitors were 35% less likely to die from COVID-19 compared to patients with hypertension not taking RAAS inhibitors (pooled RR= 0.65, 95% Confidence Intervals (CI): 0.45-0.94). To explore the association of COVID-19 and specific classes of RAAS inhibitors, we conducted a subgroup analysis of ARBs and ACEi separately from studies that provided them. Pooled risk ratio estimates from ARBs and ACEi showed a lower but not significant risk of death from COVID-19 (RR=0.93, 95% CI: 0.70-1.22) and ACEi (RR=0.65, 95% CI: 0.32-1.30).

**Conclusions:** In this meta-analysis, it was discovered that taking RAAS inhibitors, significantly decreased the risk of COVID-19 mortality in patients with hypertension. This indicates a potential protective role that RAAS-inhibitors may have in COVID-19 patients with hypertension.

**Key Questions:** *What is already known about this subject?:* Severe acute respiratory syndrome coronavirus 2 (SARS-CoV-2), responsible for the recent coronavirus disease 2019 (COVID-19) pandemic, interfaces with the renin-angiotensin-aldosterone system (RAAS) through angiotensin-converting enzyme 2 (ACE2). Recent studies have questioned whether RAAS inhibitors are safe in patients with COVID-19. However, observational studies involving patients hospitalized with COVID-19 that report the association of RAAS-inhibitors and COVID-19 severity or death have yielded conflicting findings.

*What does this study add?:* This systematic review and meta-analysis discovered that RAAS inhibitors reduced the risk of mortality by 35% in patients with hypertension hospitalized for COVID-19 (RR= 0.65, 95% CI 0.45-0.94).

*How might this impact clinical practice?:* Patients taking RAAS-inhibitors to manage their hypertension should continue to do as per current treatment guidelines.

## Introduction

Severe acute respiratory syndrome coronavirus 2 (SARS-CoV-2), responsible for the recent coronavirus disease 2019 (COVID-19) pandemic, interfaces with the renin-angiotensin-aldosterone system (RAAS) through angiotensin-converting enzyme 2 (ACE2).^1^ The current hypotheses related to the influence ACE2 may have in facilitating virus severity and mortality have been inconclusive. The increased expression of ACE2 is thought to potentially catalyze infection with COVID-19, and therefore increase the severity and risk of death.^2^ On the contrary, it has been found that ACE2 may be protective against acute lung injury.^3^

ACE2 is an 805-amino-acid, homologous to the human angiotensin-converting enzyme (ACE), with 40% identity and 61% similarity.^4^ The SARS-Cov2 virus binds to the ACE2 receptor for cell entry.^1^ Prior research has suggested that ACE-inhibitors (ACEi) and angiotensin-II blockers (ARBs), which are commonly used in patients with hypertension or diabetes, may raise ACE2 levels and thus could increase the risk of severe COVID-19 infection.^5^ Although ACE and ACE2 are two different enzymes with two different active sites, there are reports that ACE inhibitors affect the expression of ACE2 in the heart and kidneys.^6^ ARBs alter ACE2 expression, both at the mRNA and protein level.^6 7^ ACE2 is upregulated in both the renal vasculature tissue and cardiac tissue as a result of RAAS inhibitor exposure. ^8^

Individuals with cardiovascular disease including hypertension are susceptible to SARS-CoV-2 infection,^9^ and majority depend on RAAS inhibitors for hypertension control, the potential influence of ACEi and ARB during SARS-CoV-2 infection requires urgent exploration for a clarification. Despite these theoretical uncertainties regarding whether pharmacologic regulation of ACE2 may influence the infectivity of SARS-CoV-2, there is clear potential for harm related to the withdrawal of RAAS-inhibitors in patients in otherwise stable condition.

To date, observational studies involving patients hospitalized with COVID-19 that report the association of RAAS-inhibitors and COVID-19 severity or death have yielded conflicting findings.

Some studies are finding potential harmful associations of exposure to ACEi or ARBs with an increased risk of severity in COVID-19^10^, and other studies failed to confirm such findings regarding a potential harmful association.^11 12^ Many individuals with hypertension take ACE inhibitors and ARBs, but the association of RAAS-inhibitors and mortality in COVID-19 patients has not been systematically reviewed using a large number of studies. The aim of this systematic review and meta-analysis is to delineate the association between use of RAAS-inhibitors and mortality in patients with COVID-19.

We hypothesize that RAAS-inhibitors may increase mortality rates from the novel coronavirus that causes COVID-19.

## Methods

### Search Strategy and Study Selection

The authors state that all supporting data are available within the article and its online-only data supplement. We explored PubMed (MEDLINE) Scopus, the OVID databases, SCOPUS, Cochrane Library databases and medrxiv.org, using search criteria provided in the supplemental material (**Supplemental Text 1**). We invoked the Meta-analyses of Observational Studies in Epidemiology (MOOSE) during our search (**Supplemental Table 1**). We included all studies published from January 1, 2020 to May 20, 2020 that reported on the use of RAAS inhibitors (ACEi or ARBs) in patients hospitalized with COVID-19. We identified papers reporting the mortality rate in patients with and without exposure to RAAS-inhibitors. The following Medical Subject Heading (MeSH) and key words were used for the literature search of PubMed and other databases: ““receptors, angiotensin” OR “angiotensin” OR “angiotensin receptors” OR “angiotensin converting enzyme inhibitors” “Renin Angiotensin Aldosterone System” OR “Angiotensin Receptor Blocker” OR “Ace Inhibitor” OR “Angiotensin Converting Enzyme Inhibitor” AND “SARS-CoV-2” OR “COVID-19” OR “Coronavirus”. Two reviewers (ESH and AES) initially screened the titles and abstracts of all papers for eligibility. We included articles that reported the rates of death in COVID-19 patients with and without taking RAAS inhibitors. No language limitation was identified. We excluded studies that were not conducted in humans and did not report the rates of death.

### Quality Assessment and Data Extraction

Two reviewers (ESH and AES) then screened full-text articles. A third reviewer (PS) was recruited in the event of a discrepancy. Data extracted included the author, year of publication, country, sample size, the number of patients in the RAAS inhibitor group that did or did not die, and the RR/OR/HR of death in the RAAS inhibitor group compared to the non-RAAS inhibitor group. Two reviewers (ESH and AES) independently assessed the quality of the included studies. The Newcastle-Ottawa Scale (NOS) was utilized for the quality assessment of included studies. NOS scale rates observational studies based on 3 parameters: selection, comparability between the exposed and unexposed groups, and exposure/outcome assessment. This scale assigns a maximum of 4 stars for selection, 2 stars for comparability, and 3 stars for exposure/outcome assessment. Studies with less than 5 stars were considered low quality, studies receiving 5 through 7 stars were considered moderate quality, and those receiving more than 7 stars were classified as high quality.

### Data Analysis

The primary outcome of interest was the risk of mortality in patients with hypertension hospitalized for COVID-19. The exposure of interest was the use of RAAS inhibitors. Subgroup analysis of ACEi and ARB separately were conducted. We utilized the reported RR, HR, OR as the measures of the association between exposure to RAAS-inhibitors and the risk of mortality in COVID-19. For studies without measures of associations, we applied a generalized linear mixed model to calculate the odds ratios using the number of events and the sample size of each study group.^13^ Because the outcome of mortality was relatively rare, we combined RRs and HRs with ORs in the present meta-analysis and reported the pooled effect size as RRs as common risk estimates for all studies. We invoked a random-effects models to pool study results for the association between exposure to RAAS-inhibitors and the risk of mortality.^14^ We constructed forest plots to display pooled estimates. We assessed inter-study heterogeneity using *I*^2^ statistics, expressed as % (low (25%), moderate (50%), and high (75%)) and Cochrane’s *Q* statistic (significance level < 0.05) ^15 16^. Assessment of potential ascertainment bias (as might be caused by publication bias) was conducted with funnel plots, by plotting the study effect size against standard errors of the effect size, and Egger’s test. [12] We performed all statistical analyses with R software, version 3.4.3 (R, College Station, TX).

## Results

As shown in **Figure 1**, we identified a total of 337 studies from the five databases. We identified 152 studies as duplicates and excluded, leaving 185 studies. When screening titles and abstracts, we excluded 56 studies and another 129 based on full text, which left us with 2,065,805 patients from 15 studies for qualitative analysis and 61,268 patients from 11 studies for the quantitative analysis (**Table 1**).^10-12 17-27^

**Figure 1:**
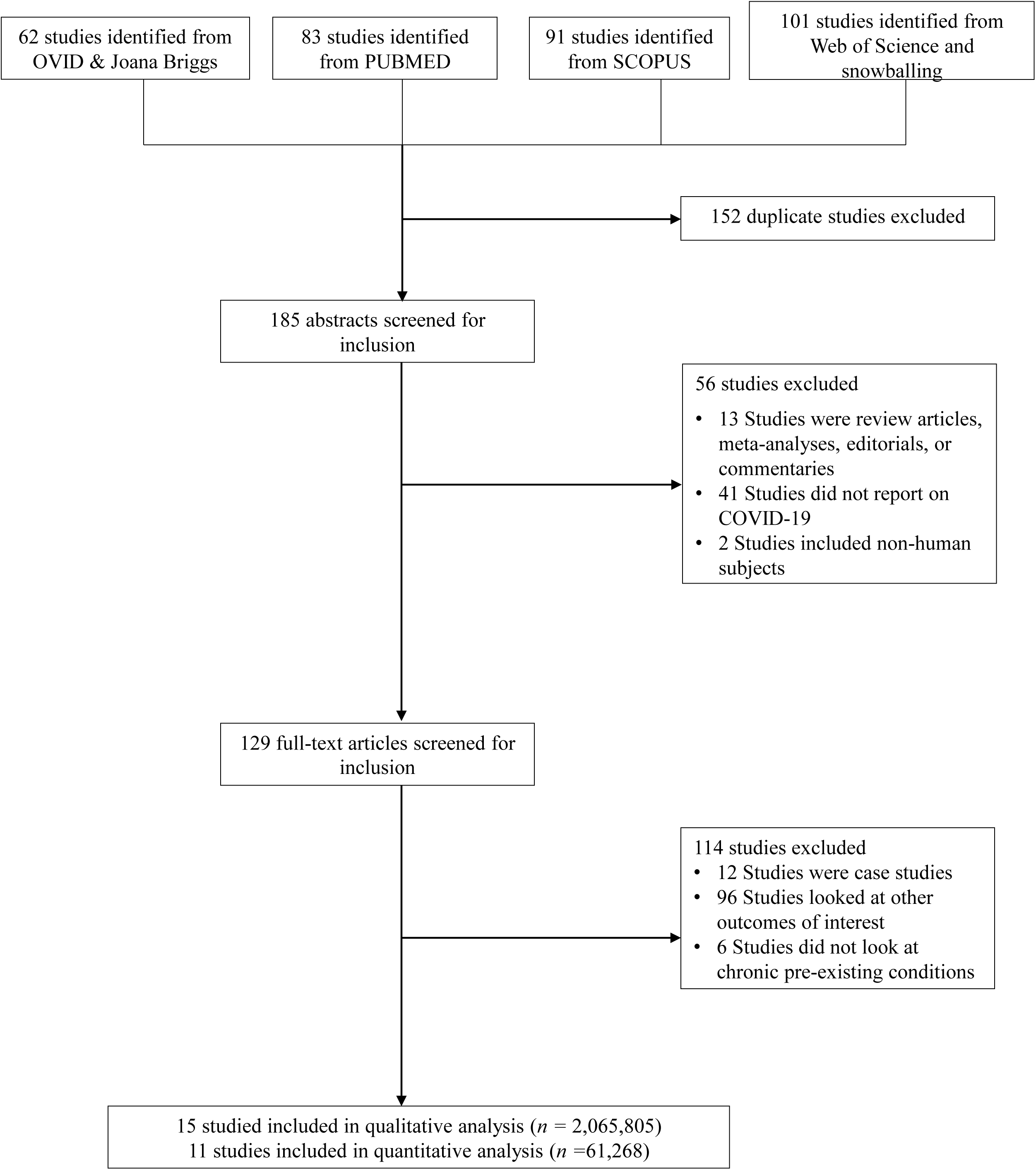
Flow Diagram.

**Table 1:**
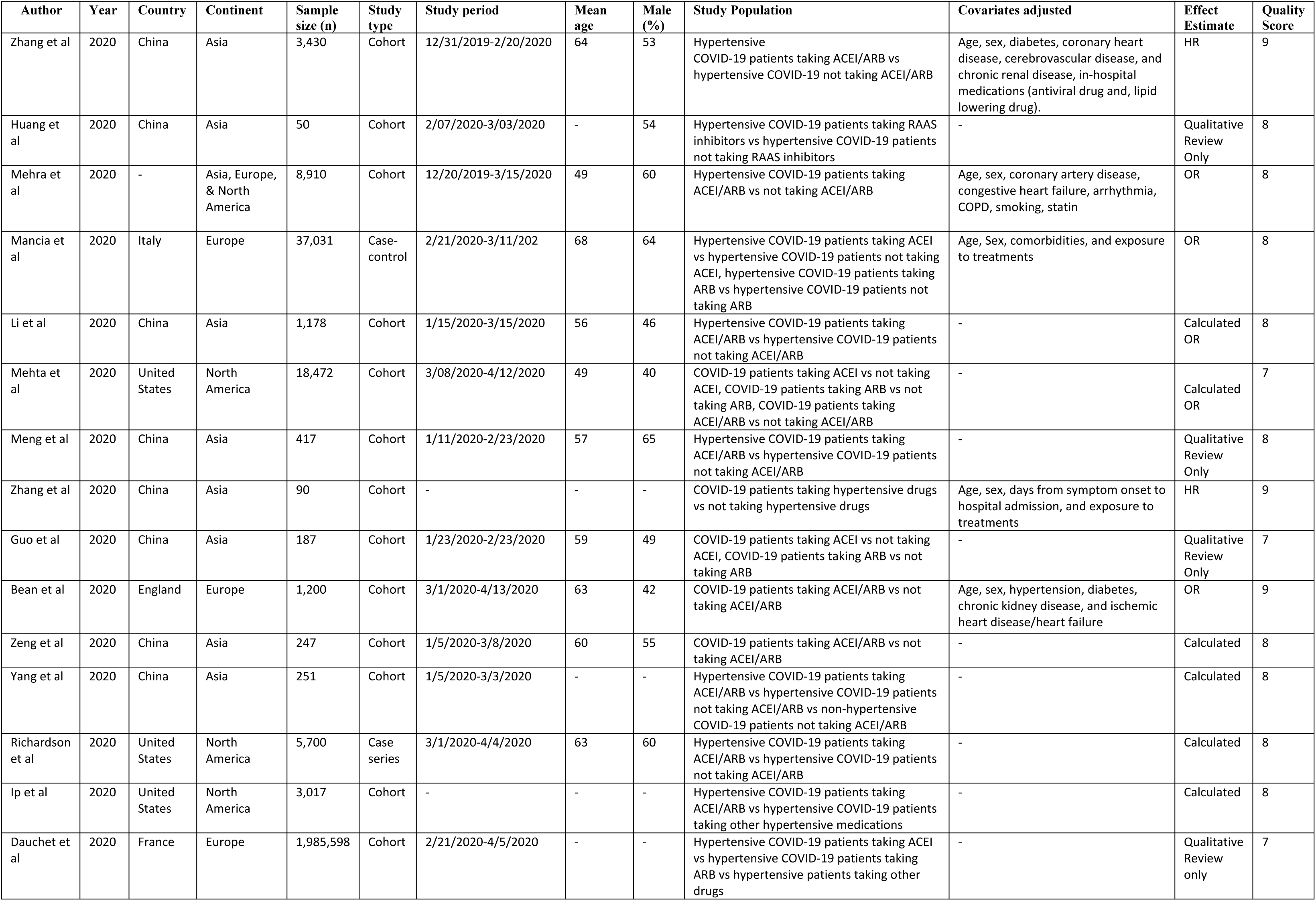
Studies of inclusion

### The association between taking RAAS inhibitors and COVID-19 mortality

Of the seven studies included in the meta-analysis exploring the association between COVID-19 mortality and RAAS exposure in patients with hypertension, three reported a significantly lower risk with mortality (**Figure 2**). No studies reported a significantly higher risk of mortality. The overall pooled estimates showed a 35% reduction in the risk of mortality (RR=0.65, 95% CI: 0.45-0.94). The inverse association between RAAS use and low risk of mortality from COVID-19, may indicate a potential protective role for RAAS inhibitors among COVID-19 patients with hypertension. Between-study heterogeneity was high (*I*^2^= 80, p<0.01).

**Figure 2.**
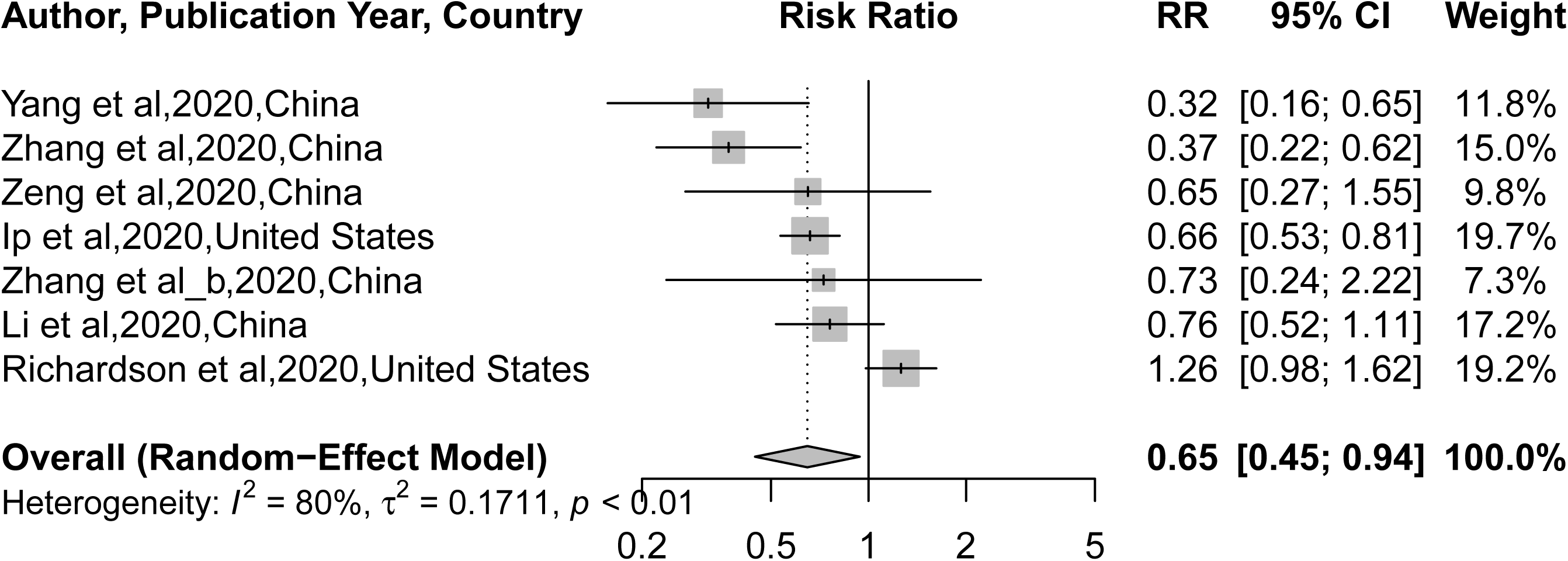

### Subgroup analysis of the association of ARBs and ACEi with mortality from COVID-19

To explore the association of COVID-19 and specific classes of RAAS inhibitors, we conducted a subgroup analysis of ARBs and ACEi separately from studies that provided them. Three studies provided risk ratios estimates (or data to calculate the RR) for ACEi (**Figure 3)** and three for ARBs (**Figure 4**). Pooled risk ratio estimates from ARBs and ACEi showed a low risk of death from COVID-19 (RR=0.93, 95% CI: 0.70-1.22) and ACEi (RR=0.65, 95% CI: 0.32-1.30). However, the association was not significant. Between-study heterogeneity was moderate (I^2^= 58, p=0.09) from ARBs and high (I^2^= 90, p<0.01) for ACEi.

**Figure 3.**
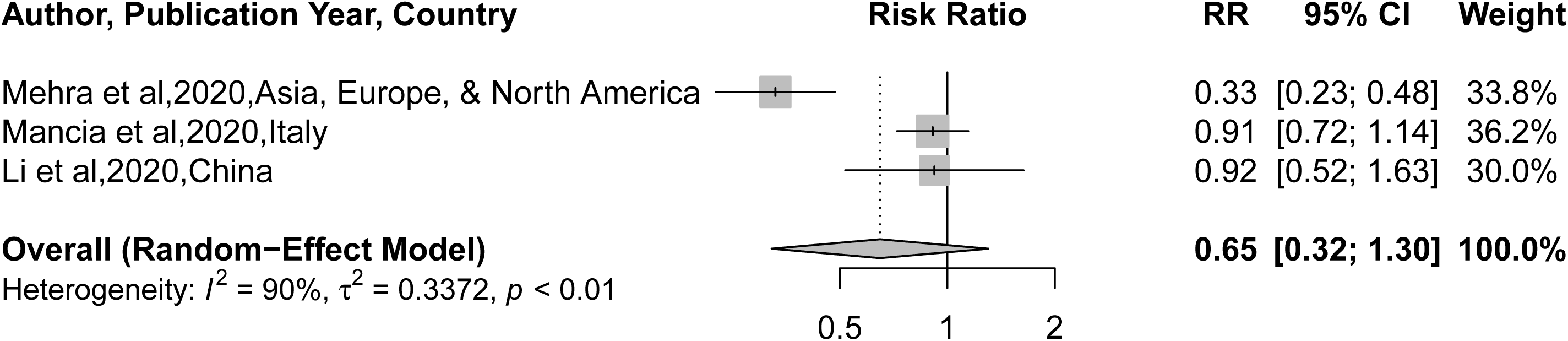

**Figure 4.**
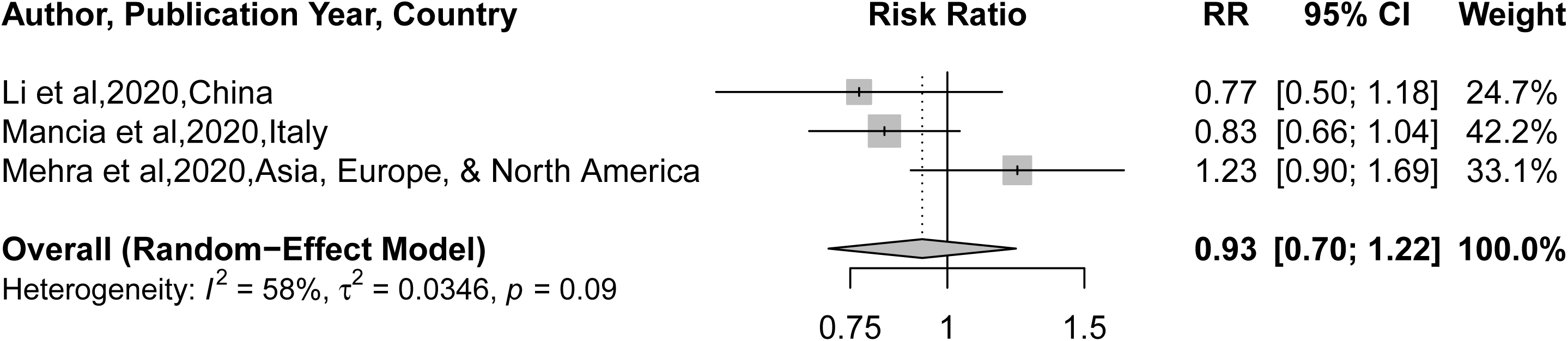

### Sensitivity Analyses, Publication Bias Publication and Study Quality

We conducted two types of sensitivity analyses. First, limiting our analysis to the 7 RAAS inhibitor studies whose sample size consisted of only individuals with hypertension, we excluded and replaced one study at a time from the meta-analysis and calculating the pooled RR for the remaining studies. No substantial changes from pooled RR were observed when other studies were removed in turn. The pooled RR ranged from 0.57 to 0.72 (p<0.0001 for all) (**Supplementary Figure 1**). Second, by including all studies (main analysis plus 3 studies with sample size not limited to population with hypertension, see **table 1**), pooled RR from sensitivity analysis ranged from 0.68 to 0.82 (p<0.0001 for all) (**Supplementary Figure 2**). We did not find significant funnel plot asymmetry for the Egger test for RAAS inhibitor, ACEi and ARBs (p > .05). Upon visual inspection, the funnel plots appeared symmetrical for RAAS inhibitors, ACEi and ARBs (**Supplemental Figures 3-6**). The median study quality score was 8 out of 9 (range=7-9).

## Discussion

This systematic review and meta-analysis of cohort studies consisting of 2,065,805 patients with a global representation suggests that the treatment of hypertension with RAAS-inhibitors is associated with a lower risk of mortality in patients with COVID-19. This finding is important, for the association between RAAS-inhibitor exposure and mortality in COVID-19 patients has been inconclusive thus far. This topic has been heavily debated, and some studies even have interpolated a risk of taking RAAS-inhibitors using data from previous coronavirus outbreaks and preclinical studies. [6]

RAAS-inhibitors have been found to mitigate the risk of severe lung injury by reducing the activation of the RAAS through the inactivation of angiotensin II^3^ and the generation of angiotensin-(1-9) ^4^ and angiotensin-(1-7) ^28^. Angiotensin-(1-7) binds to the G protein-coupled receptors Mas to mediate various physiological effects including vasorelaxation, cardio protection, anti-oxidation and inhibition of angiotensin II -induced signaling. This is one hypothesized mechanism illustrating how the treatment of chronic conditions with RAAS-inhibitors may be beneficial in COVID-19 patients.

Alternatively, it is hypothesized that the biological mechanisms of RAAS inhibitors may predispose COVID-19 patients to severe disease and mortality. This was explained by the observation that SARS-CoV-2 enters a cell by binding to the membrane-bound ACE2 receptors. Animal models suggest that ACEis and ARBs increase membrane-bound ACE2 receptors, to which then increases the availability of cells for SARS-CoV-2 to bind and cellular entry.^6^ This hypothesis has sparked a debate in populations, for many individuals taking RAAS inhibitors have grown concerned that their medications may be predisposing them to developing COVID-19, and later dying from it.^29^ Our meta-analysis and systematic review support the notion that RAAS inhibitor exposure does *not* increase COVID-19-related mortality but rather shows beneficial effect.

### Strengths and Limitations

Limitations of our study include possible selection bias in the published literature as a result of the strict COVID-19 testing algorithm employed in the early stages of the pandemic. This may have resulted in missed COVID-19 cases or deaths. Nevertheless, this is the largest quantitative synthesis of evidence on the association between RAAS-inhibitor exposure and COVID-19 mortality. The regions with the highest burden of COVID-19, including Asia, Europe, and North America, were represented thus increasing the external validity of our findings. The sample size included in this study was also quite large, allowing us to thoroughly cover a large population.

## Conclusion

This systematic review and meta-analysis found that COVID-19 patients with hypertension who take RAAS-inhibitors are protected from COVID-19 mortality compared to hypertensive patients not taking RAAS-inhibitors. Patients taking RAAS-inhibitors to manage their chronic diseases may continue to do as per current treatment guidelines and based on the clinical judgment of their health care providers.

## Data Availability

All data is available within the manuscript.

## Contributorship

AS, PS, ES, and VC conceived the study. AS, EH, and PS conducted the literature search. AS and PS completed data analysis. AS, DL, PD, VC, AL JO, EH and PH interpreted the data. AS, ES, and PS wrote the manuscript. All Authors agreed to the manuscript in its final form

## Acknowledgements

We would like to acknowledge Melissa Butt for reviewing and proving helpful feedback.

## Patient and Public Involvement

Patients or the public were not involved in the preparation or dissemination of this manuscript

## Ethics Statement

This is a systematic review and meta-analysis and individual patient was not used. Therefore we did not need IRB or an ethics board approval.

## Data Sharing Statement

All data relevant to the study are included in the article or uploaded as supplementary information

## Supplementary material for Ssentongo et., al 2020

**<u>Supplemental text 1: PubMed Search Terms</u>**

((((((“receptors, angiotensin”[MeSH Terms] OR “angiotensin”[All Fields])) OR “angiotensin receptors”[All Fields]) OR (“angiotensin”[All Fields] OR “(((((“angiotensin converting enzyme inhibitors”[Pharmacological Action] OR “angiotensin-converting enzyme inhibitors”[MeSH Terms]) OR (“angiotensin converting enzyme inhibitors”[All Fields]) OR OR “ace inhibitor”[All Fields])) OR “angiotensin-converting enzyme inhibitors”[MeSH Terms]) OR angiotensin converting enzyme inhibitors”[All Fields])

**Supplemental Table 1:** Meta-Analyses and Systematic Reviews of Observational Studies (MOOSE)

|  | Topic | Page number |
| --- | --- | --- |
| **Title** | Identify the study as a meta-analysis (or systematic review) | 1 |
| **Abstract** | Use the journal's structured format | 2 |
| **Introduction** | **Present:** | 4 |
|  | The clinical problem | 4 |
|  | The hypothesis | 4 |
|  | A statement of objectives that includes the study population, the condition of interest, the exposure or intervention, and the outcome(s) considered | 4 |
| **Sources** | **Describe:** |  |
|  | Qualifications of searchers (eg, librarians and investigators) | 5 |
|  | Search strategy, including time period included in the synthesis and keywords | 5 |
|  | Effort to include all available studies, including contact with authors | 5 |
|  | Databases and registries searched | 5 |
|  | Search software used, name and version, including special features used (e.g. explosion) | 5-6 |
|  | Use of hand searching (e.g, reference lists of obtained articles) | 5 |
|  | List of citations located and those excluded, including justification | 5 |
|  | Method of addressing articles published in languages other than English | 5 |
|  | Method of handling abstracts and unpublished studies | 5 |
|  | Description of any contact with authors | 5 |
| **Study Selection** | **Describe** |  |
|  | Types of study designs considered | 5 |
|  | Relevance or appropriateness of studies gathered for assessing the hypothesis to be tested | 5 |
|  | Rationale for the selection and coding of data (eg, sound clinical principles or convenience) | 5 |
|  | Documentation of how data were classified and coded (eg, multiple raters, blinding, and inter-rater reliability) | 5 |
|  | Assessment of confounding (e.g. comparability of cases and controls in studies where appropriate) | 5-6 |
|  | Assessment of study quality, including blinding of quality assessors; stratification or regression on possible predictors of study results | 5-6 |
|  | Assessment of heterogeneity | 6 |
|  | Statistical methods (eg, complete description of fixed or random effects models, justification of whether the chosen models account for predictors of study results, dose-response models, or cumulative meta-analysis) in sufficient detail to be replicated | 6 |
| **Results** | **Present** | 6-7 |
|  | A graph summarizing individual study estimates and the overall estimate | 15 |
|  | A table giving descriptive information for each included study | Table 1 |
|  | Results of sensitivity testing (eg, subgroup analysis) | 6-7 |
|  | Indication of statistical uncertainty of findings | 6-7 |
| **Discussion** | **Discuss** | 8-9 |
|  | Strengths and weaknesses | 8-9 |
|  | Potential biases in the review process (eg, publication bias) | 8-9 |

\*Modified from Stroup DF, Berlin JA, Morton SC, Olkin I, Williamson GD, Rennie D, et al. Meta-analysis of observational studies in epidemiology: a proposal for reporting. Meta-analysis Of Observational Studies in Epidemiology (MOOSE) group. JAMA 2000;283:2008–12. Copyrighted © 2000, American Medical Association. All rights reserved.

**Supplementary Figure 1.**
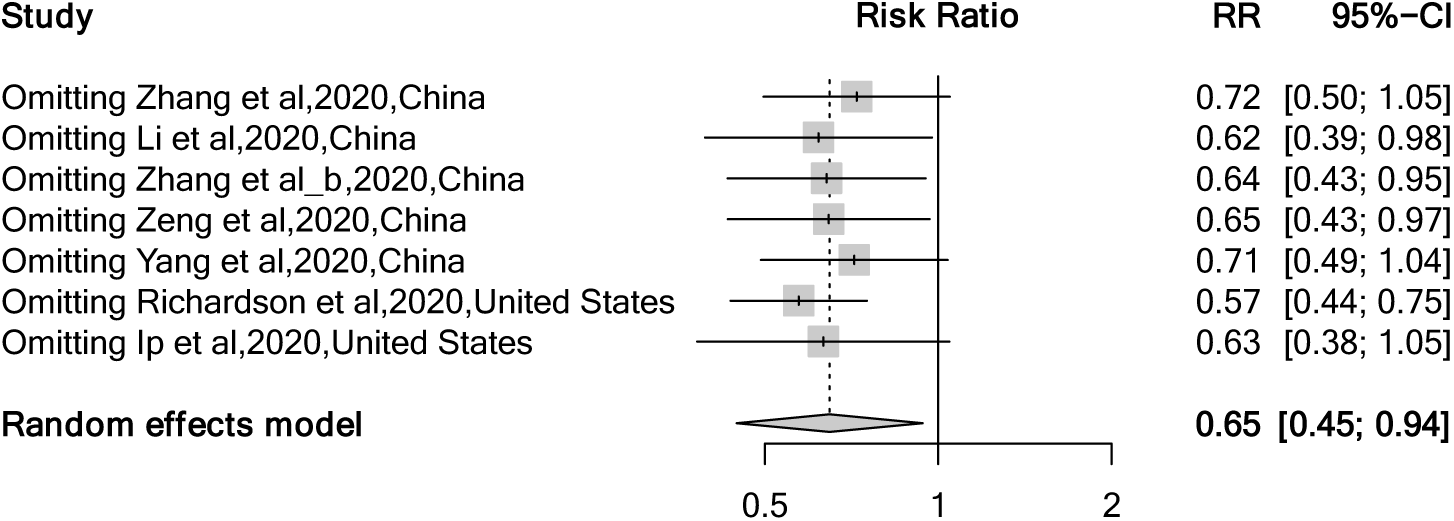
Sensitivity analysis of random-effects meta-analysis of studies evaluating the association of ACEi/ARB with mortality in patients with hypertension hospitalized with COVID-19 (only study population hypertension)

**Supplementary Figure 2.**
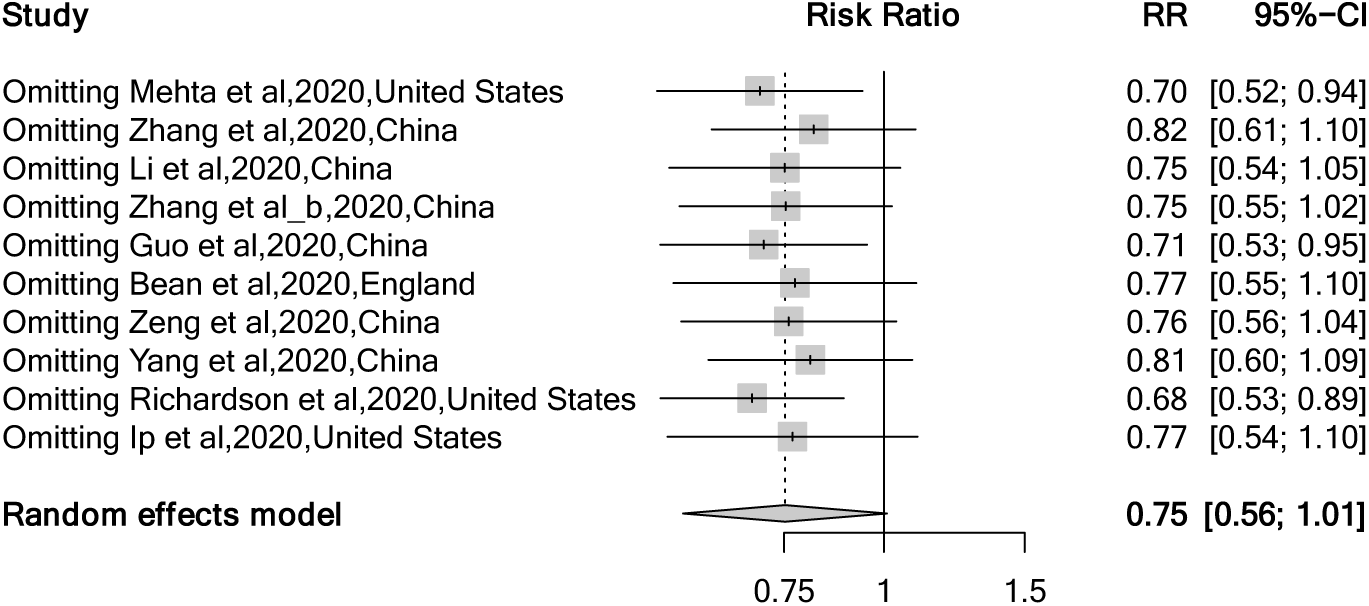
Sensitivity analysis of random-effects meta-analysis of studies evaluating the association of ACEi/ARB with mortality in patients with hypertension hospitalized with COVID-19 (study population with and without hypertension)

**Supplementary Figure 3.**
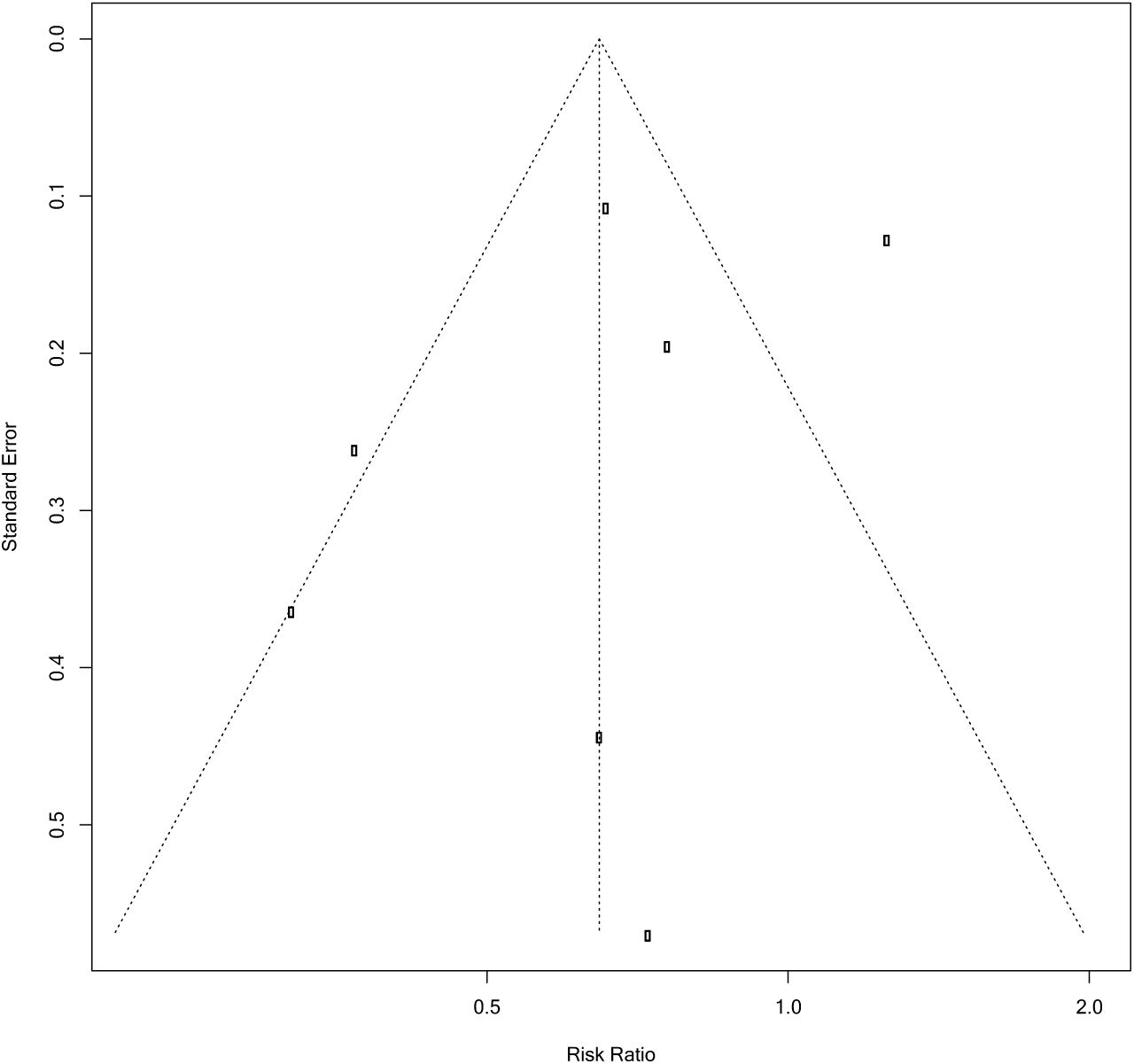
Funnel plot depicting publication bias for studies evaluating the association of ACEi/ARB with mortality in patients with hypertension hospitalized with COVID-19 (only study population hypertension)

**Supplementary Figure 4.**
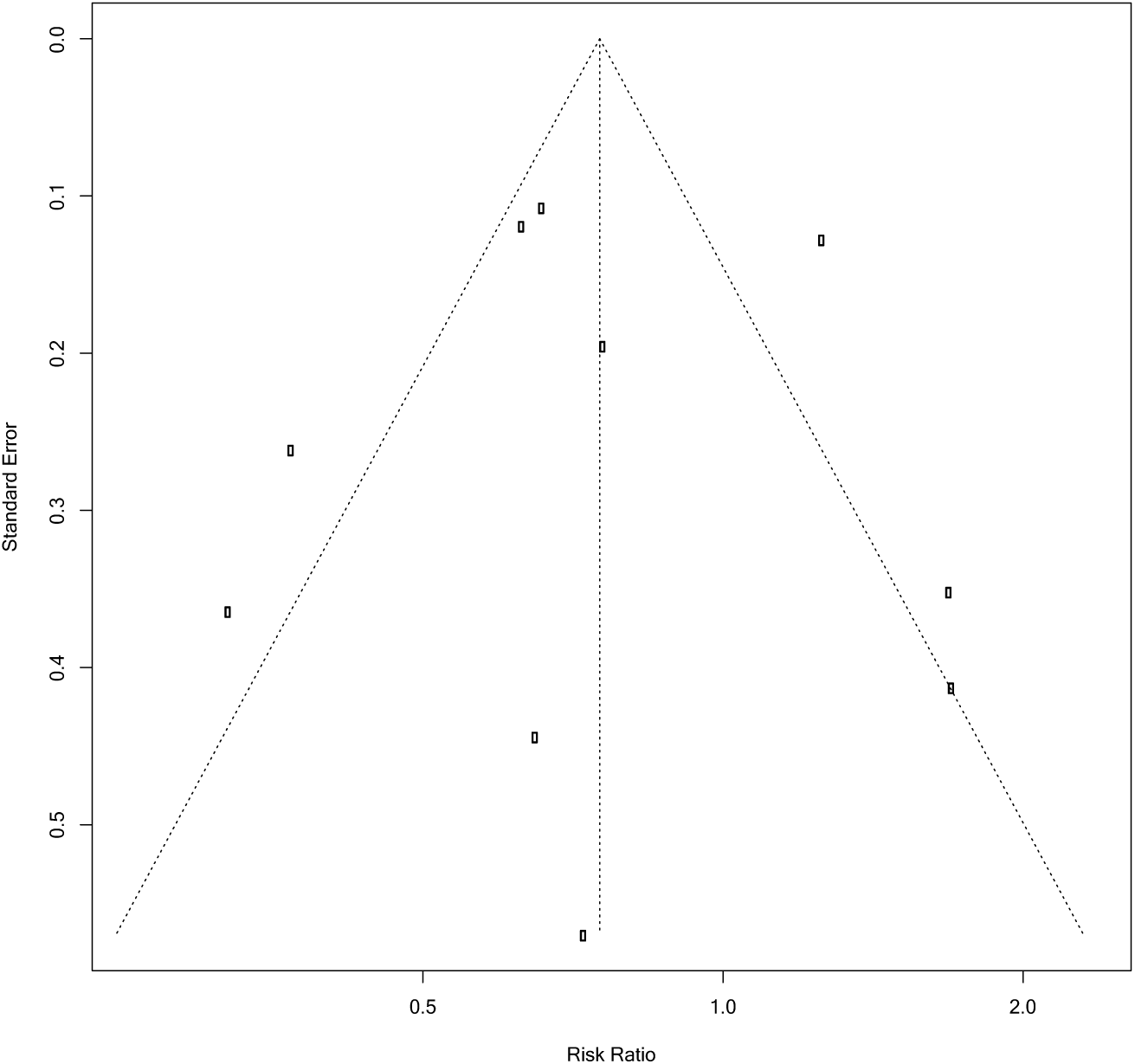
Funnel plot depicting publication bias for studies evaluating the association of ACEi/ARB with mortality in patients with hypertension hospitalized with COVID-19 (study population with and without hypertension)

**Supplementary Figure 5.**
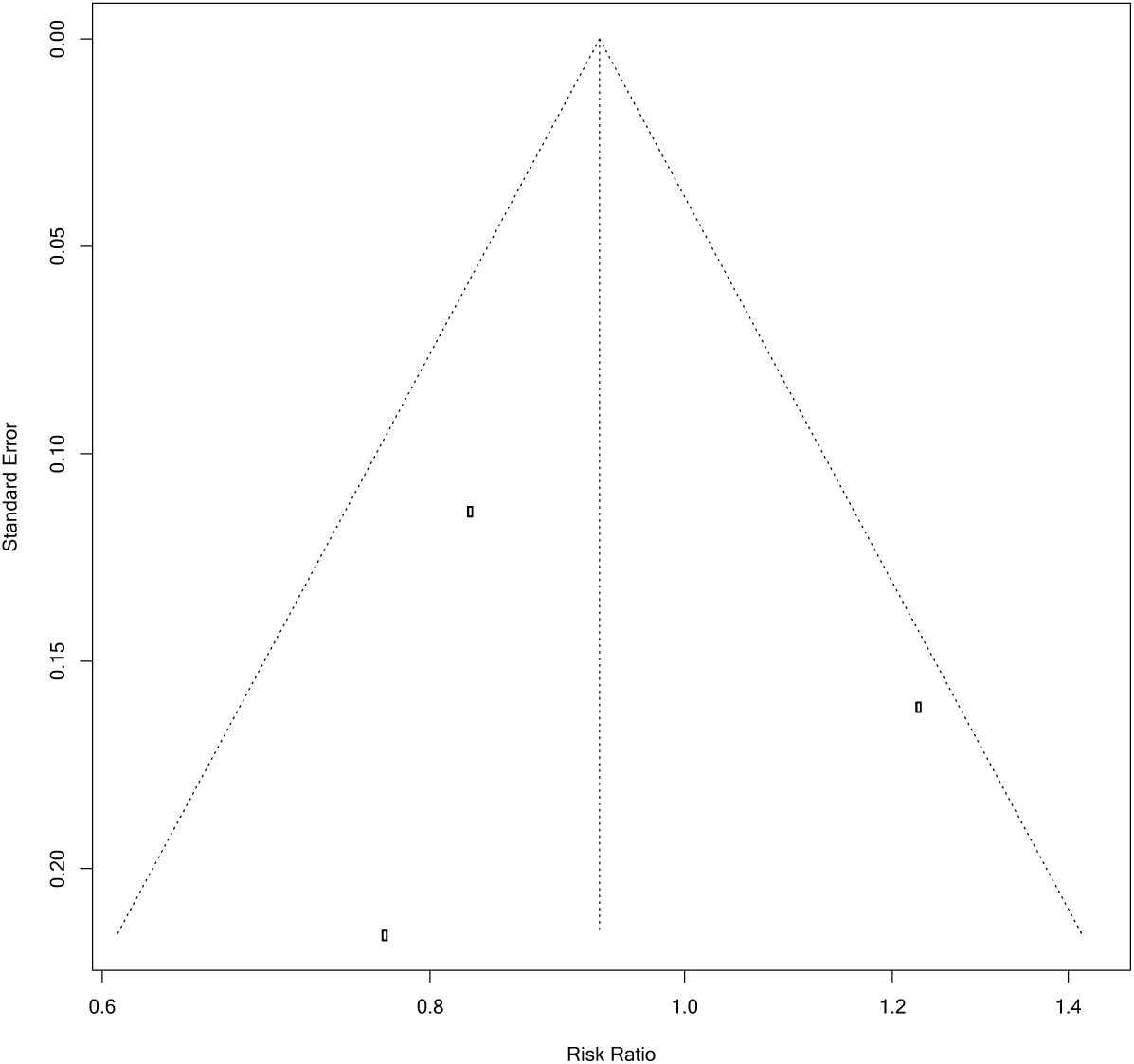
Funnel plot depicting publication bias for studies evaluating the association of ARB with mortality in patients with hypertension hospitalized with COVID-19.

**Supplementary Figure 6.**
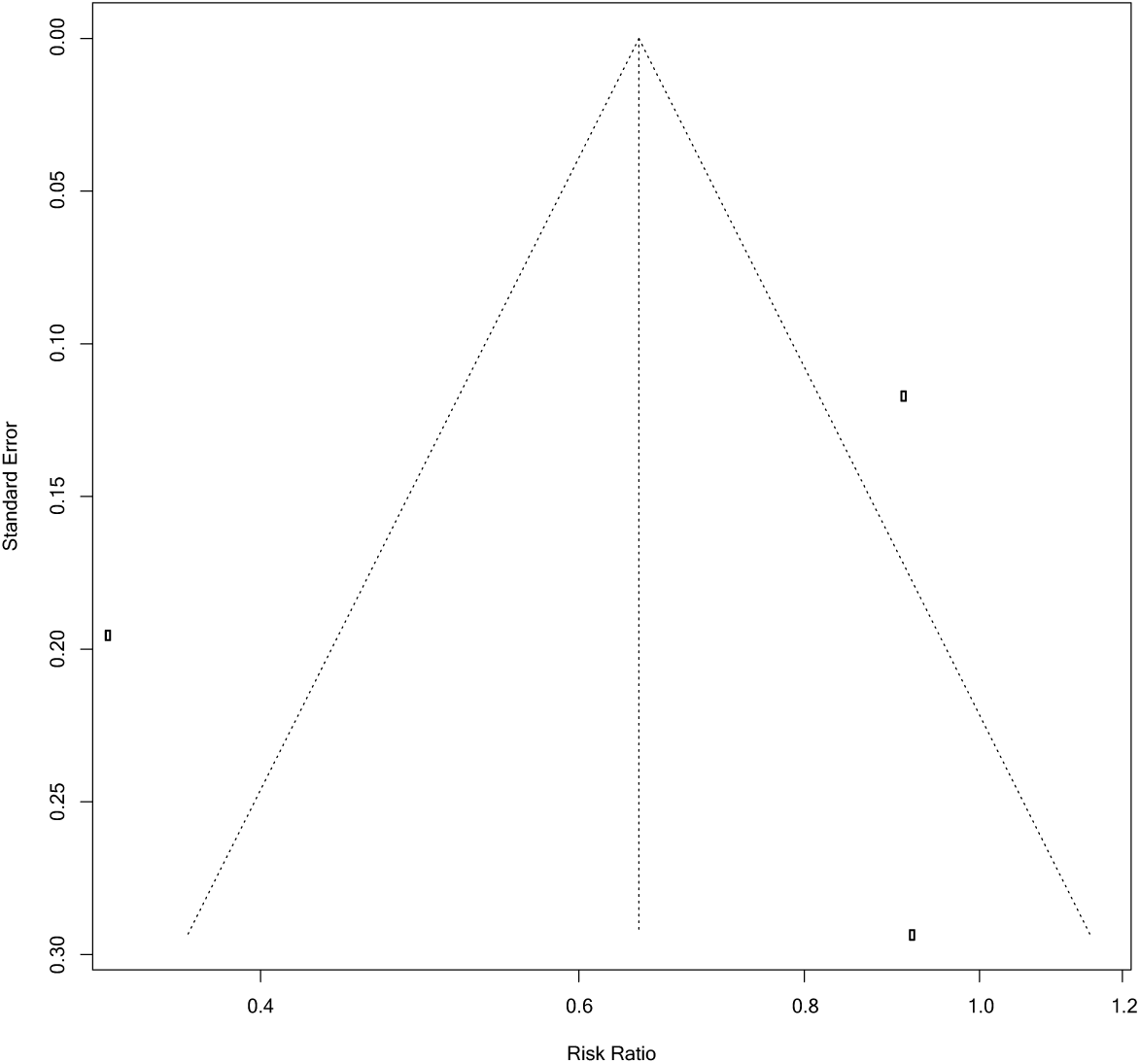
Funnel plot depicting publication bias for studies evaluating the association of ACEi with mortality in patients with hypertension hospitalized with COVID-19.

